# Computational Fluid Dynamics Based Risk-Stratification of Modified Blalock-Taussig-Thomas Shunt Thrombogenicity

**DOI:** 10.1101/2025.05.03.25326898

**Authors:** Yi Qiao, Ethan Penn, Jacob Miller, Scott Bugenhagen, Ram Rohatgi, Kelsey Mercer, Blaire Kulp, Jinli Wang, Pirooz Eghtesady, Guy M. Genin, Edon J Rabinowitz, David Bark

**Author notes:** Corresponding author: David Bark, PhD, Associate Professor, Pediatric Hematology Oncology Associate Professor, Biomedical Engineering, Associate Professor, Division of Biology and Biomedical Sciences, https://barkmechanics.com, 5103 McDonnell Pediatric Research Building | Washington University School of Medicine in St. Louis | St. Louis, MO 63108; Edon J Rabinowitz MD, Assistant Professor, Pediatric Critical Care & Cardiology Washington University in St. Louis 660 S. Euclid Ave, Campus Box 8116 St. Louis, MO 63110. **COI:** none. IRB: This study was approved by the Washington University in St Louis Institutional Review Board (IRB ID# 202303162). **CENTRAL MESSAGE:** Computational modeling reveals that modified Blalock-Taussig-Thomas-Shunt geometry directly alters blood flow biomechanics linked to thrombosis, guiding surgical strategies to reduce clotting risk. **PERSEPECTIVE STATEMENT:** This study lays the groundwork for precision anticoagulation by integrating patient-specific vascular anatomy, fluid biomechanics, and physics to inform engineering-based surgical planning. Independent of drug-based strategies, this approach offers a novel, mechanistically distinct path to thrombosis prevention in cardiovascular surgery. **CENTRAL PICTURE LEGEND:** CFD enables thrombosis risk-stratification based on shunt geometry and flow dynamics. (co-senior author) (Edon J Rabinowitz, David Bark).

## Abstract

**Background:** The modified Blalock-Taussig-Thomas shunt (mBTTS) is a critical palliative procedure for infants with single-ventricle physiology, but thrombosis-related occlusion affects 8-12% of cases and carries nearly 50% mortality. Meanwhile, existing antithrombotic strategies fail to address the hemodynamic factors driving thrombosis, highlighting the need for a deeper understanding of flow dynamics in shunt failure.

**Objectives:** This study aims to identify how mBTTS geometry influences hemodynamics and thrombosis risk, providing quantitative guidance for surgical planning and shunt design optimization.

**Methods:** We used patient-specific imaging data to construct 54 idealized mBTTS configurations, systematically varying key geometric factors; pulmonary artery diameter, shunt diameter, and insertion angle. Using computational fluid dynamics, we analyzed how these variables influence wall shear rate (WSR), elongational strain rate (ESR), and turbulence intensity (TI); hemodynamic parameters known to affect thrombosis risk, to identify patterns linked to thrombosis.

**Results:** We computationally identified optimal geometric configurations. Peak WSR and ESR were primarily located at bifurcation points, while peak TI was concentrated within the shunt channel. Shunt insertion distal to the right carotid artery with a 60° insertion angle and with a 4.0mm shunt graft demonstrated the most favorable hemodynamic profiles to prevent clots. Statistical analysis confirmed strong correlations between geometric parameters and flow characteristics.

**Conclusion:** Results provide a framework for optimizing mBTTS design to reduce thrombosis risk based on hemodynamic risk factors, including actionable recommendations for shunt placement and design. These insights provide a foundation for hemodynamically guided surgical interventions with potential to improve survival rates in this high-risk patient population and for broader applications in cardiovascular surgery.

## INTRODUCTION

The modified Blalock-Taussig-Thomas shunt (mBTTS, **Fig. 1a**) is a life-saving palliative procedure for infants with cyanotic or single ventricle heart disease. However, thrombosis occurs in 8–12% of cases and carries up to 50% mortality^1–7^. Survivors of early complications also face a significantly lower 5-year survival (33–40% vs. 77%)^1,8,9^. These outcomes suggest that either shunt geometry promotes thrombosis or certain patients have biological risk factors. Current therapies target biochemical pathways with limited efficacy, but neglect flow-driven triggers^10–13^. We propose a precision health approach to optimize mBTTS geometry and reduce thrombogenic flow in this high-risk population.

**Figure 1:**
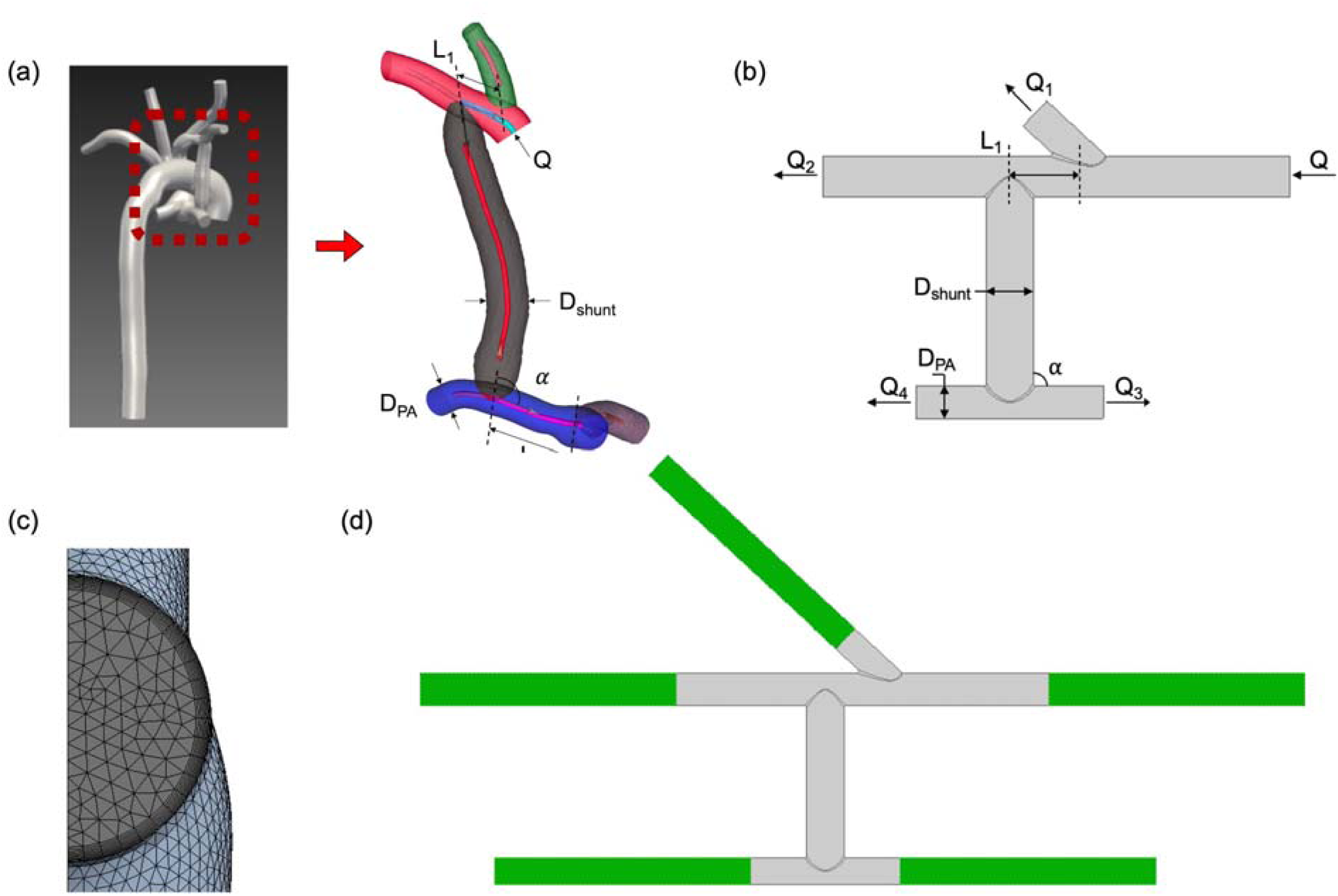
Development and Characterization of the Idealized mBTTS Computational Model. **(a)** Patient-derived mBTTS geometry showing key parameters: *L*_1_ (distance to right carotid), *D_shunt_* (shunt diameter), *D_PA_* (pulmonary artery diameter), and α (shunt-PA angle). **(b)** Simplified “H-model” preserving critical anatomy while standardizing vessel paths and bifurcation for controlled geometric analysis. Boundary conditions include a constant inlet velocity (Q); outlet flows (Q_1_–Q_4_) follow Murray’s Law. **(c)** Cross-sectional mesh with 10 inflation layers (first layer: 1×10^-^□ m) for boundary layer resolution. **(d)** Full domain with inlet/outlet extensions (10× subclavian diameter) to ensure developed flow; extensions shown in darker shade (green online).

To explore the role of geometry in shunt thrombosis, we retrospectively reviewed mBTTS placements in infants at Washington University (2018–2023)^14^. Despite a limited sample size, trends in geometric differences between patent and occluded shunts suggest that targeted adjustments may help prevent thrombosis.

To investigate the role of geometry, we used computational fluid dynamics (CFD) to identify shunt configurations that minimize thrombogenic flow. We systematically varied geometry to assess its impact on three key hemodynamic factors: wall shear rate (WSR), elongational strain rate (ESR), and turbulence intensity (TI). WSR quantifies the velocity gradient at the vessel wall, reflecting the shear stress exerted by the fluid. ESR describes the rate of flow deformation along the velocity direction. In contrast to WSR, where adjacent layers of blood slide past one another, ESR involves blood elements being stretched or elongated along the direction of flow, and is a parameter known to extend long chain molecules^15^. High WSR/ESR can promote thrombosis by activating platelets, through a von Willebrand factor (VWF)-dependent mechanism, where fluid stress is understood to extend VWF, exposing key sites (**Fig. 2**)^11,16–23^. TI quantifies velocity fluctuations due to turbulence. Higher TI amplifies platelet-platelet and platelet-protein interactions and mechanical stress, contributing to thrombosis risk^24,25^.

**Figure 2:**
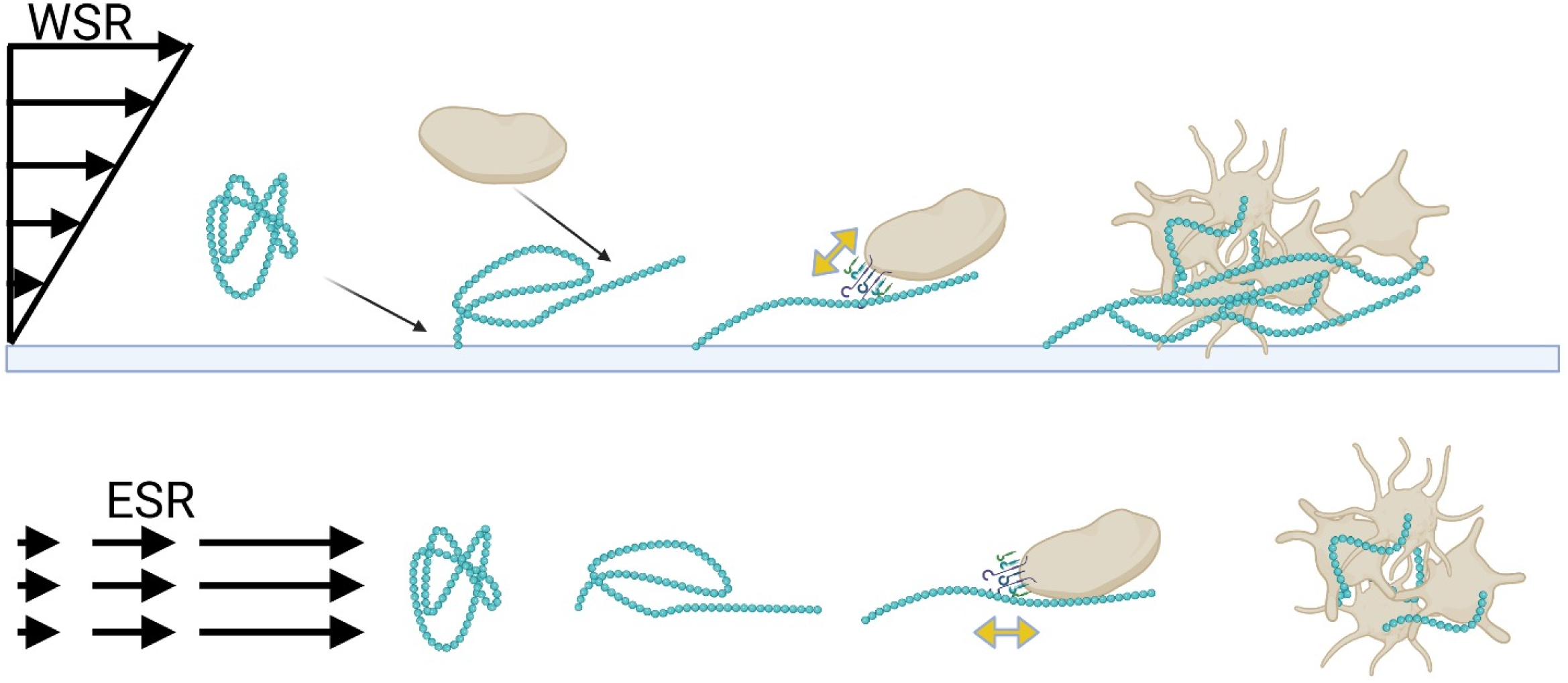
Effects of Wall Shear Rate (WSR) and Elongational Strain Rate (ESR) on Platelet Activation. (Top) High WSR elongates von Willebrand factor (VWF), promoting platelet adhesion and aggregation via GPIb-IX-V binding. (Bottom) ESR stretches VWF along flow lines, exposing platelet binding sites and enhancing VWF self-association. Both high WSR and ESR drive VWF-mediated thrombosis, while low shear may promote clotting via prolonged cell residence and coagulation activation.

Our simulations enable systematic exploration of geometric configurations difficult to study surgically, providing precise control of key variables. By analyzing how geometry affects WSR, ESR, and TI, we aim to identify design principles that reduce thrombosis risk and guide surgical planning.

## METHODS

### Study Overview and Model Development

Building on prior identified geometric risk factors for mBTTS thrombosis^14^, we developed idealized computational models to systematically assess how specific geometric features influence thrombogenic flow. This study was approved by the Washington University IRB (ID# 202303162).

Using SimVascular (Palo Alto, CA), we first created 3D models of mBTTS anatomy from patient CT/MR imaging. Analysis of these models informed the development of a simplified “H” model (**Fig. 1**) that enabled precise control of key geometric parameters while intentionally excluding confounding factors, e.g. vessel tortuosity. This approach follows established methodology in computational hemodynamics, where idealized models are used to isolate fundamental relationships between flow and geometry before reintroducing complexity, the primary goal of this study^26–31^. Inlet and outlet extensions (10× subclavian vessel diameter) ensured fully developed flow conditions at the mBTTS region (**Fig. 1d**). Future validation studies will reintroduce patient-specific anatomy to confirm clinical relevance in more complex geometries.

### Geometric Parameter Selection and Model Generation

Four key geometric parameters identified from our previous clinical study^14^. Three parameters were evaluated across the minimum, median, and maximum values observed in our clinical cohort (**Table 1**) that can be studied in the idealized H model: the distance between shunt insertion and right carotid artery (RCA) insertion (L_1_, −7 to +7 mm, with negative values indicating insertion distal to the RCA), the angle between the shunt and pulmonary artery (*α*, 60° to 120°), and the pulmonary artery diameter (D_PA_, 2.5 to 5.0 mm). The fourth parameter, shunt diameter (D_shunt_), was evaluated at two clinically standard sizes: 3.5 mm and 4.0 mm. This resulted in 54 unique geometric configurations, each maintaining consistent vessel paths while varying only the specified parameters.

**Table 1.** Geometric Parameter Ranges Used in Computational Analysis of mBTTS Configurations. Summary of parameters across 54 modeled configurations. L_1_ is the distance from shunt to right carotid artery (negative=proximal, positive=distal). α is the shunt-PA insertion angle. D_PA_ and D_shunt_ are pulmonary artery and shunt diameters, respectively. Values were derived from patient-specific imaging data^14^.

| | $L_1$, mm | $\alpha$, ° | $D_{PA}$, mm | $D_{shunt}$, mm |
| --- | --- | --- | --- | --- |
| Minimum | -7 | 60 | 2.5 | 3.5 |
| Median | 0 | 90 | 3.75 |  |
| Maximum | 7 | 120 | 5.0 | 4.0 |

Outflow ratios were calculated using Murray’s Law,^32^ with vessel radii derived from our patient cohort’s median values (**Table E1**). These ratios determined initial flow distribution among the right carotid artery, right subclavian artery, and both pulmonary arteries. While this approach provides physiologically reasonable flow splitting based on vessel diameter relationships, it simplifies physiology. Pulmonary vascular resistance (PVR) relative to systemic vascular resistance (SVR) determines actual flow distribution in clinical settings. Our model assumed uniform outlet pressure, effectively equalizing downstream resistance across vascular beds. This allowed isolation of geometric effects on flow, while acknowledging that patient-specific PVR/SVR ratios would alter flow distributions in vivo. Future models incorporating patient-specific resistances could improve the translational value, especially for pre-surgical planning in patients with pulmonary hypertension or altered vascular resistance.

### Mesh Generation and Validation

Surface and volume meshes were generated using ANSYS Fluent (Canonsburg, PA). To accurately capture boundary layer effects, ten prismatic inflation layers were added near vessel walls with tetrahedral elements throughout the remaining volume. The inflation layers used a first-layer thickness of 1×10⁻L m and a growth rate of 1.1, achieving a wall y-plus value of 0.5 (<1) to ensure well-resolved near-wall flow characteristics (**Fig. 1c**). Mesh convergence analysis balanced computational efficiency with solution accuracy. Element counts ranged from 7,500 to 1,437,000, with convergence achieved at ∼476,300 elements. Models were standardized to use approximately 800,000 cells each, with minor adjustments for anatomical variations. Mesh evaluated using standard quality metrics including skewness, orthogonal quality, and aspect ratio. Quality thresholds were set to ensure numerical reliability: maximum skewness below 0.82, minimum orthogonal quality above 0.20, and maximum aspect ratio below 40.

### Computational Flow Analysis

Blood flow simulations were performed using ANSYS Fluent (Canonsburg, Pennsylvania) with the implicit, steady-state, k-ω SST turbulence model and low Reynolds number correction. Blood was treated as a Newtonian fluid, consistent with blood behavior at relatively high shear, with constant density (1060 kg/m³) and viscosity (0.004 kg/m·s). We applied no-slip boundary conditions at rigid, impermeable walls and specified constant velocity inlet profiles based on median flow velocities measured by Doppler echocardiography in our patient cohort^14^. Steady-state flow was used to minimize confounding variables and isolate geometric effects. While the inlet Reynolds number (1830) indicated laminar flow for a straight vessel, we employed the k-ω SST model with low-Reynolds correction to accurately capture potential flow transitions and near-wall effects in complex bifurcation regions.

For each configuration, we calculated three key hemodynamic parameters associated with thrombosis risk. The first was wall shear rate (WSR), which was calculated as:

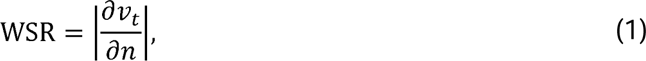

where *v_t_* is blood velocity parallel to the wall and *n* is the distance perpendicular to the wall. The second was elongational strain rate, ESR, which was quantified as the rate of stretching along the flow streamlines:

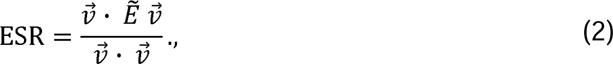

where 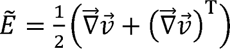 is the strain rate tensor and 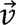 is the blood velocity at a point.

The third was the turbulence intensity, TI, which was quantified as:

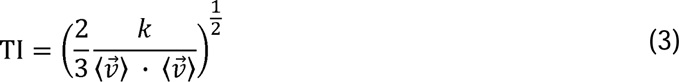

where 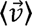 is the time average of 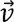 and *k* is the turbulent kinetic energy from the Reynolds-averaged Navier-Stokes model used in the current work. This parameter represents the amount that velocity fluctuates, i.e. the turbulence level, relative to the average velocity.

Finally, to classify the flow based on whether it stretches blood components (extensional) or rotates them (rotational), we computed a flow-type parameter, *λ*:^33^

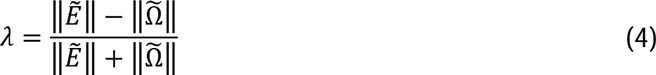

where 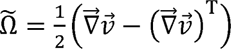 is the vorticity tensor, and 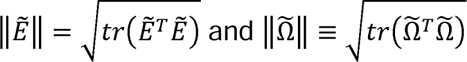 are Frobenius norms. *λ* = 1 corresponds to pure extensional flow, *λ* = −1 represents pure rotational flow, and *λ =* 0 indicates simple shear.

### Optimization

To identify optimal geometric configurations, we applied a composite score that balances ESR, WSR, and TI:

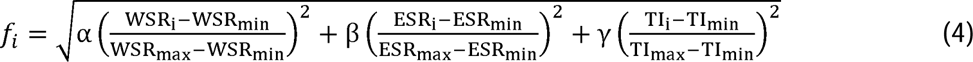

Where the subscript *max* and *min* represent the extrema over all permutations of geometric parameters, the subscript *i* indicates a specific combination. The weighting factors *α,β*, and *γ* are determined using Principal Component Analysis (PCA) method, which quantifies each variable’s contribution based on data variance. Derived from the Euclidean distance formulation, the equation normalizes each variable to prevent bias and enables fair comparison. The function *f* objectively evaluates different geometric configurations. A lower f-value indicates a more optimal configuration, while a higher f-value reflects less favorable flow characteristics.

### Statistics

Data were summarized using frequencies with percentages, means with standard deviations, or medians with interquartile range, as appropriate. Spearman’s correlation coefficient (r) was calculated to examine the association between parameters and shunt insertion outcomes. Differences among the parameters with 3 levels were compared using an analysis of variance (ANOVA) model. When the overall model was significant, Tukey-adjusted pairwise comparisons were performed with significant comparisons reported. If the normality assumption was violated, rank-transformed data were analyzed. For parameters with two levels, either the two-sample t-test or the Wilcoxon rank-sum test was used, depending on the distribution of the data. Analyses were conducted using SAS version 9.4 (SAS Institute Inc, Cary, NC), and P values < 0.05 were considered significant.

## RESULTS

### Spatial Distribution of Peak Flow Characteristics

Analysis of the 54 mBTTS configurations revealed distinct peak locations for each flow metric (**Fig. 3c, f, i, Table E3**). WSR peaks occurred primarily at bifurcation points, with 46.3% at the distal attachment corner of the shunt-subclavian artery junction, 29.6% at the proximal attachment corner, 20.4% (11 models) at the right carotid artery insertion, and 3.7% (2 models) along the shunt wall. ESR peaks were similarly concentrated at junctions, with 66.7% occurring at the distal corner of the shunt-subclavian artery junction, 27.8% at the right carotid artery bifurcation, and 1.8% at the shunt-pulmonary artery junction. TI peaks differed, with 70.4% occurring within the shunt conduit, 11.1% at the inlet, 7.4% distal to the shunt insertion, and 5.6% split equally between the carotid and shunt-pulmonary artery junctions. Representative contour plots show the distribution of WSR, ESR, and TI across the model (**Fig. 3a, b, d, e, g, h**).

**Figure 3:**
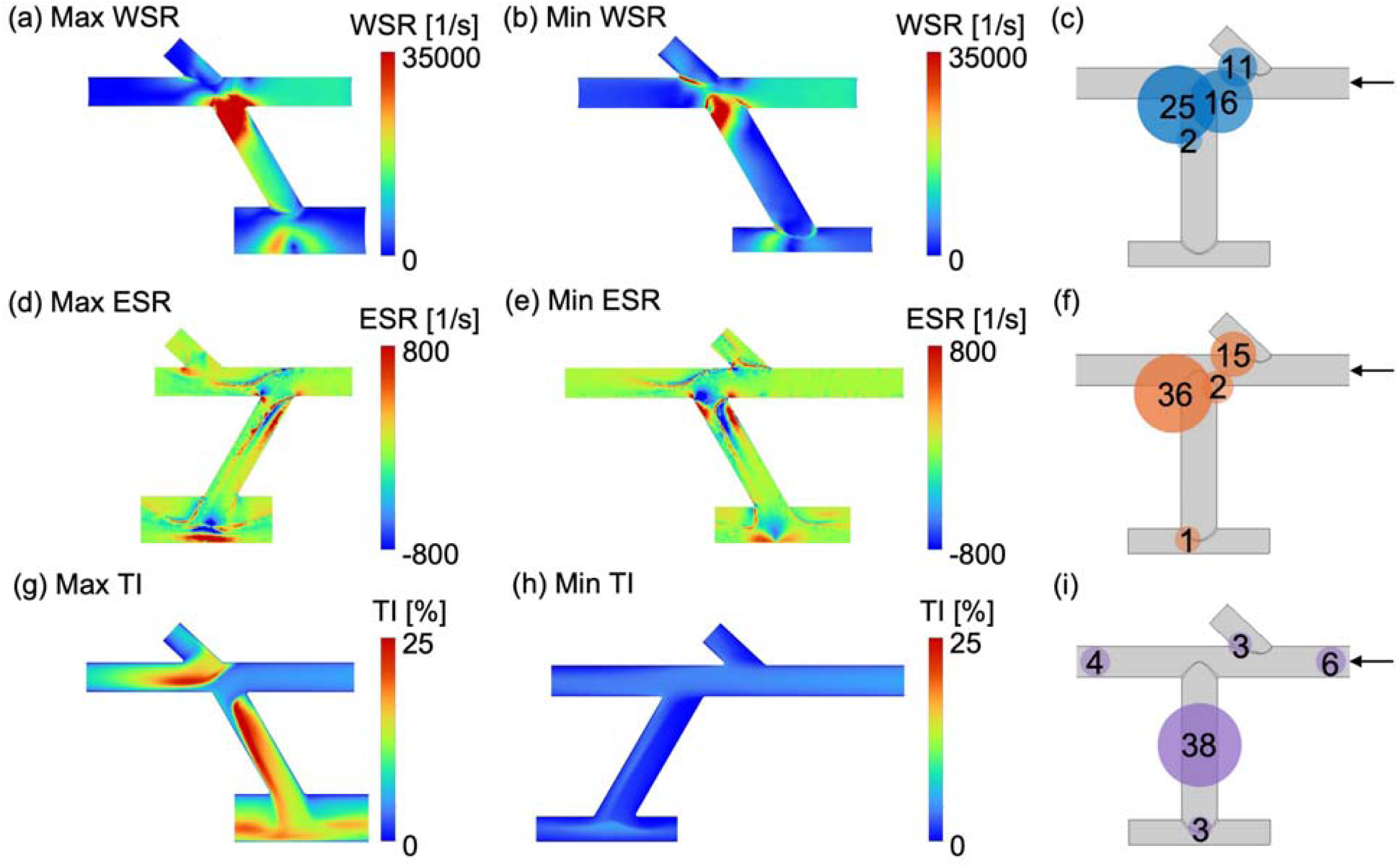
Spatial Distribution and Comparison of Extreme Flow Parameters Across mBTTS Geometries. Contour plots show models with maximum and optimized values for each hemodynamic parameter over a cardiac cycle. Schematics summarize peak locations across 54 configurations; circle size reflects frequency of peak occurrence. **(a)** Maximum WSR: 61404s^−^, (*D_PA_*=5.0mm, *D_shunt_* =3.5mm). **(b)** Optimized WSR: 24216s^−^ (*D_PA_* =2.5mm, *D_shunt_* =4.0mm). **(c)** WSR peaks (blue): mostly at shunt-subclavian anastomosis **(d)** Maximum ESR: 7911s^−^ (*L*_1_=-7mm, *α*=60°). **(e)** Optimized ESR: 2741s^−^ (*L*_1_=7mm, *α*=120°). **(f)** ESR peaks (orange): at distal shunt-subclavian junction (66.7%) and carotid bifurcation (27.8%). **(g)** Maximum TI: 33.9%, (*D_PA_*=5.0mm, *D_shunt_* =3.5mm). **(h)** Optimized TI: 6.8% (*L*_1_=7mm, *α*=60°). **(i)** TI peaks (purple): primarily within the shunt (70.4%). Key anatomical features labeled: right carotid artery (RCA), right subclavian artery (RSCA), shunt, and pulmonary arteries (PA). Arrows indicate flow direction. Percentages based on all simulated configurations.

### Influence of Geometric Parameters on Flow Characteristics

Statistical analysis demonstrated clear correlations between geometric parameters and flow characteristics (**Fig. E1, Table E2**). Moving the shunt insertion from proximal (L_1_ = −7 mm) to distal (L_1_ = 7 mm) moderately decreased ESR and TI, with WSR showing a similar but nonsignificant trend (r = −0.26, p = 0.054). A larger shunt-to-PA angle (α) reduced ESR (r = - 0.40) but increased TI (r = 0.37). D_PA_ strongly correlated with all flow metrics (r > 0.3), while shunt diameter (D_shunt_) was inversely associated with WSR (r = −0.33) and showed weak, nonsignificant effects on ESR and TI.

Overall and pairwise comparisons confirmed these trends (**Fig. 4**). Significant effects were observed for α and D_PA_ (p < 0.05), with D_PA_ increasing all flow metrics, especially TI (p < 0.0001). L_1_ significantly impacted ESR (p = 0.028) and TI (p = 0.0042), while D_shunt_ selectively influenced WSR (p = 0.016). These findings identify D_PA_ and α as key determinants of all flow metrics, with L_1_ and D_shunt_ exerting more targeted effects.

**Figure 4:**
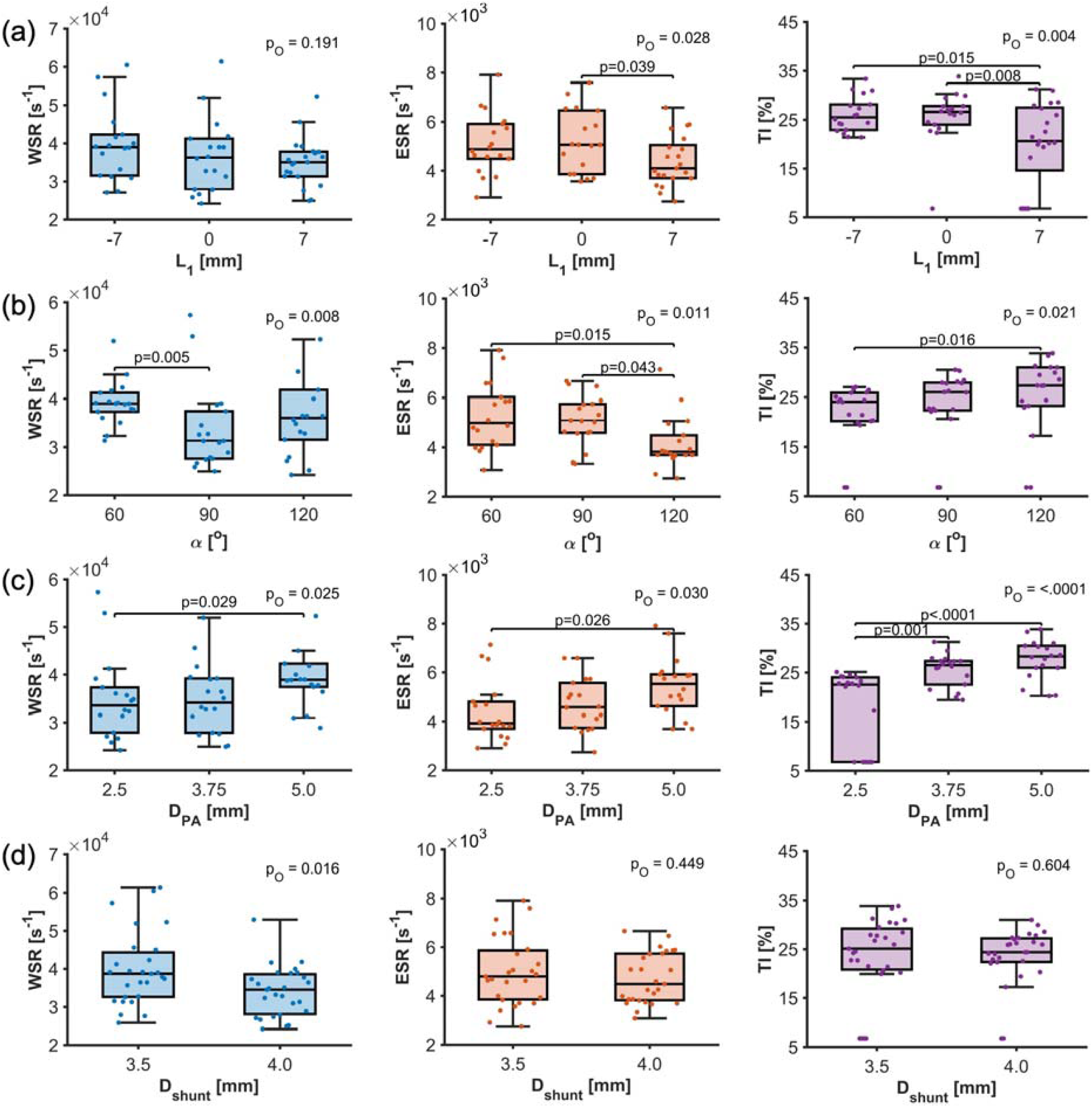
Impact of Geometric Parameters on Hemodynamic Metrics. Analysis of the relationships between geometric parameters and flow metrics (WSR, ESR, TI). Box plots display median, quartiles, and range for each parameter level. Overall (P_O_) and pairwise p-values highlight significance across all geometric variations. (a) L_l_ (b) α (c) D_PA_ (d) D_shunt_. Changes in any parameter produced significant shifts in at least one flow metric, emphasizing the sensitivity of mBTTS geometry to thrombosis-related flow alterations.

Models exhibiting extreme values for each flow metric demonstrated distinct geometric configurations (**Table 2**). Maximum WSR (61400 s^-1^) occurred in a model with D_PA_= 5.0 mm and D_shunt_ = 3.5 mm, while minimum WSR (24200 s^-1^) was observed with D_PA_ = 2.5 mm and D_shunt_ = 4.0 mm. Peak ESR (7910 s^-1^) was found in a configuration with L_1_= −7 mm and α = 60°, while minimum ESR (2740 s^-1^) occurred with L_1_ = 7 mm and α = 120°. Maximum TI (33.9%) was observed in the same configuration as peak WSR, while minimum TI (6.8%) occurred in a model with L_1_= 7 mm, α = 60°, D_PA_ = 2.5 mm, and D_shunt_ = 4.0 mm.

**Table 2:** Geometric Parameters of Models Exhibiting Extreme Flow Characteristic. Configuration details and peak values for models demonstrating maximum and optimized values of each flow characteristic (Wall Shear Rate, WSR; Elongational Shear Rate, ESR; and Turbulence Intensity, TI). Each row represents a specific model configuration identified during systematic analysis of 54 geometric variants.

| | Model | $L_1$, mm | $\alpha$, ° | $D_{PA}$, mm | $D_{shunt}$, mm | Peak Value |
| --- | --- | --- | --- | --- | --- | --- |
| Max peak WSR | DP 44 | 0 | 120 | 5.0 | 3.5 | 61400 |
| Min peak WSR | DP 11 | 0 | 120 | 2.5 | 4.0 | 24200 |
| Max peak ESR | DP 36 | -7 | 60 | 5.0 | 3.5 | 7910 |
| Min peak ESR | DP 32 | 7 | 120 | 3.75 | 3.5 | 2740 |
| Max peak TI | DP 44 | 0 | 120 | 5.0 | 3.5 | 33.9% |
| Min peak TI | DP 15 | 7 | 60 | 2.5 | 4.0 | 6.8% |

### Geometric Configurations Associated with Flow Optimization

Analysis of all configurations that consistently minimized or maximized potentially adverse flow characteristics was identified through the optimization method defined by Eqn. 4, (**Fig. 5**). By applying the optimization approach, the weighting factors for WSR, ESR, and TI were determined as 0.38, 0.25, and 0.37, respectively, when calculating the optimization factor, *f* (Table E3). We ranked all 54 models by *f*-value and identified clear trends. The six models with the lowest *f*-values shared L_1_ = 7 mm and D_PA_ = 2.5 mm, while the five highest *f*-value models had D_shunt_ = 3.5 mm and D_PA_ = 5.0 mm. These patterns suggest *f* is particularly sensitive to one or more of these parameters. Surface plots of *f* against geometric parameter pairs (**Fig. E2**) further supports this. Lower f-values are observed in models with either L_1_ = 7 mm and the smallest D_PA_, or D_shunt_ = 4.0 mm and the smallest D_PA_. Together, these findings indicate that a distal shunt insertion (L_1_= 7 mm), a 4mm D_shunt_, and a smaller D_PA_ are associated with more favorable flow characteristics.

**Figure 5.**
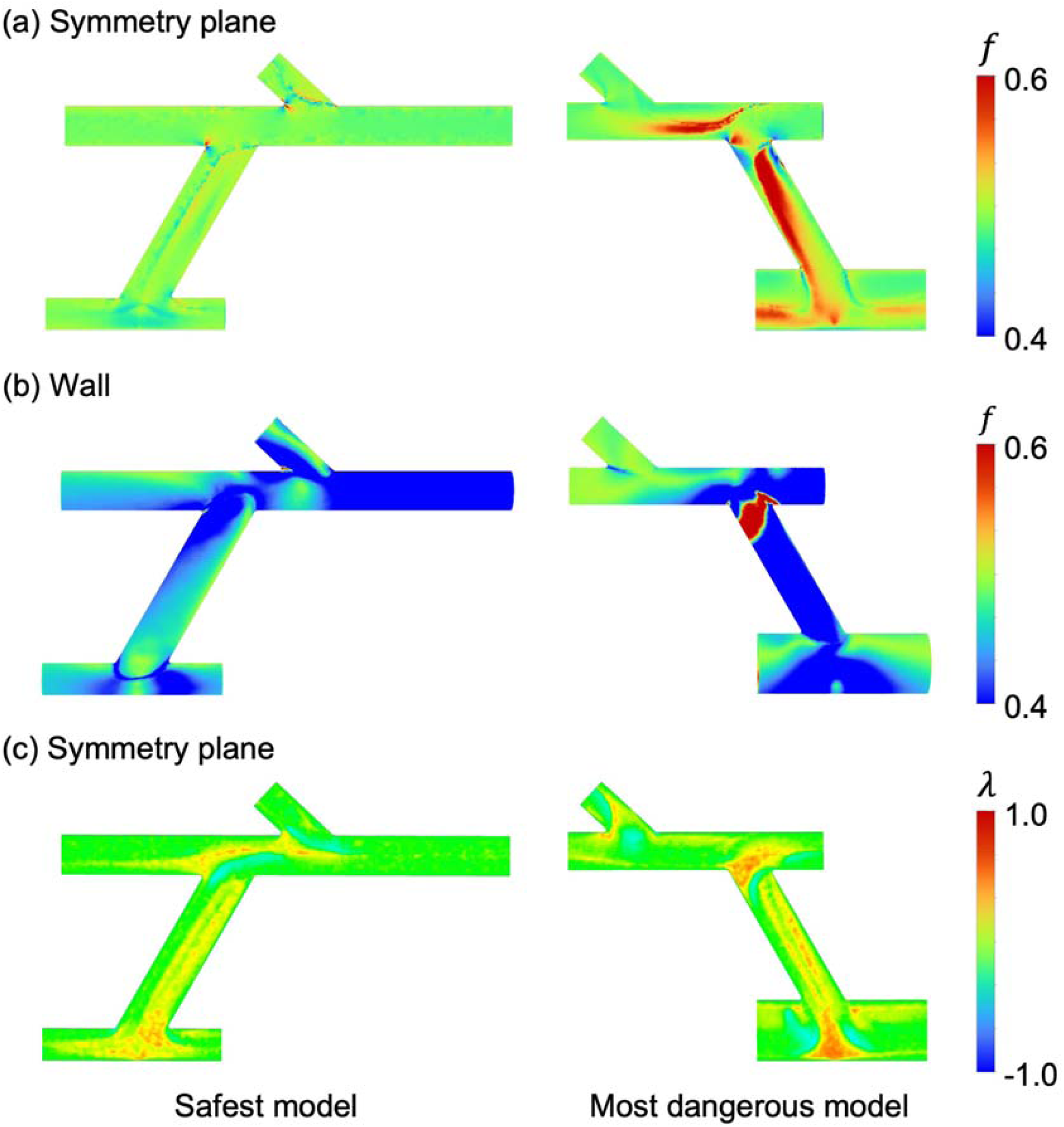
Comparative Analysis of Safest and Most Thrombogenic mBTTS configurations. Models selected based on composite optimization factor *f*, reflecting balance of WSR, ESR, and TI. The “safest” model minimizes all three metrics; the “most dangerous” maximizes them. (a–b) Contour plots of *f* on the symmetry plane and vessel wall for both models. (c) Flowtype parameter (λ) on the symmetry plane, comparing dominant flow regimes.

The configuration demonstrating the most favorable flow profile (*f* = 0.14) combines a distal shunt insertion (L_1_ = 7 mm), acute pulmonary artery angle (α = 60°), minimal pulmonary artery diameter (D_PA_ = 2.5 mm), and larger shunt diameter (D_shunt_ = 4.0 mm). This configuration produced WSR values of 32,300 s^-1^, ESR of 3080 s^-1^, and TI of 6.8%. *f* remained uniformly low across the domain (**Fig. 5a, b**). In contrast, the configuration associated with the highest-risk flow characteristics (*f* = 0.90) combined proximal shunt insertion (L_1_= −7 mm), obtuse pulmonary artery angle (α = 120°), maximal pulmonary artery diameter (D_PA_ = 5.0 mm), and smaller shunt diameter (D_shunt_ = 3.5 mm). This arrangement generated WSR values of 60,500 s^-1^, ESR of 5920 s^-1^, and TI of 33.3%. High *f* values were concentrated at the shunt-subclavian junction and shunt–PA connection, especially along the shunt and near bifurcating zones, primarily driven by TI. Intermediate geometric configurations showed consistent trends where distal shunt insertions (L_1_ >0) combined with acute angles (α = 60°) consistently produced lower *f* values, independent of vessel diameters. Models with D_PA_ ≤3.75 mm demonstrated more moderate flow characteristics across all measured parameters compared to those with larger pulmonary artery diameters.

To further evaluate local flow behavior, the λ parameter, associated with elongational flow relative to rotation and linked to VWF extension, was mapped across the symmetry plane. For the lowest *f* value configuration, the λ distribution (**Fig. 5c**) was centered around zero, corresponding to simple shear, with small positive values near bifurcations and stagnation points. Negative λ was minimal, at the shunt-subclavian junction, indicating limited rotational flow. For the greatest *f* value configuration, positive λ occurred predominantly along the shunt, at the shunt subclavian and at shunt-PA junction, aligning with elevated ESR. Negative λ remained limited, suggesting dominant extensional flow over rotational features.

## DISCUSSION

Our CFD analysis of 54 mBTTS configurations demonstrated that specific geometric features directly influence thrombosis-related flow. Distal shunt placement relative to the right carotid artery improved ESR and TI, and acute insertion angles (60°) reduced adverse flow at bifurcations. Smaller pulmonary arteries (≤3.75 mm) also dampened turbulence. Most importantly, all geometric changes led to significant shifts in WSR, ESR, and/or TI, highlighting the sensitivity of thrombogenic flow to surgical configuration.

mBTTS failure from partial or complete shunt occlusion poses a life-threatening risk requiring emergent intervention to restore patency^8,34–38^. Rapid diagnosis demands high clinical vigilance and swift action. The associated healthcare costs are substantial, estimated at $1.3–$2.9 million per patient or $74 billion for all complex CHD cases^39^. Current mBTTS antithrombotic guidelines, largely unchanged since 2013^1^, recommend acute therapies such as high-dose heparin, vasopressors, ventilatory support, and catheter-based or surgical thrombus removal, sometimes with ECMO. Preventive strategies rely on antiplatelet and anticoagulant agents but have poor evidence, limited monitoring, and risks of bleeding or heparin-induced thrombocytopenia^1,8,38,40^. Without a mechanistic understanding of mBTTS thrombosis, optimizing therapies remains difficult.

Despite evidence that mBTTS thrombosis involves VWF-dependent mechanisms not targeted by current therapies, hemodynamic-based prevention strategies are lacking, but can be arguably hemostatically safer than prescribing antithrombotics. Flow conditions known to increase thrombosis risk can be mitigated: WSR and ESR at vessel junctions and TI within the shunt. Our virtual analysis identified an optimized geometry; distal shunt insertion, acute pulmonary artery angle, small PA diameter, and large shunt diameter, that reduced peak WSR, ESR, and TI by ∼50% compared to the least favorable configuration. Several of these features are surgically modifiable, offering opportunities for evidence-based improvement. While anatomical constraints and surgical exposure may limit full implementation, these findings highlight practical targets to reduce thrombogenic flow and guide postoperative risk assessment.

### Mechanistic Implications for Thrombosis

There have been multiple advances in our understanding of mechanobiological pathways triggering thrombosis. High wall shear rates can induce shear-induced platelet activation and aggregation through VWF, known to occur as a function of exposure time and shear stresses, observed in less than a second for shear stress exceeding 400 dynes/cm^2^ (10,000 s^-1^)^41,42^, as observed at bifurcations—especially the shunt-subclavian anastomosis.

Elongational flow is increasingly understood to induce VWF extension at lower thresholds than shear-alone ^43,44^, exposing platelet binding sites. Mechanical tension across platelet GPIb-IX-V (>20 pN) through VWF triggers granule release and calcium signaling, leading to integrin activation and platelet aggregation^45^. ESR is highest within the flow core and minimal at vessel walls, making their role in shunt thrombosis unclear. VWF extension and binding with platelets within flow could prime them for wall deposition. Also, combined ESR and WSR may lead to VWF self-association, potentiating further thrombotic risk^22^. More studies are needed to fully understand the role of ESR relative to WSR in causing a thrombotic environment.

TI, which peaked within the shunt conduit, represents a third mechanical pathway to thrombosis. Turbulent flow enhances platelet-platelet collisions and exposure to repeated mechanical stress, potentially amplifying the prothrombotic effects initiated at bifurcations^17^. Multiple studies have suggested that transient changes in stress can increase the likelihood for platelet activation^20,46^. Turbulence, can also lead to a converse situation of bleeding risk due to its potential role in acquired von Willebrand syndrome (loss of high molecular weight VWF multimers)^47^. The relative role of ESR, WSR, and turbulence in thrombosis remain unclear for an mBTTS, which is why all are considered in the current study.

The optimization factor *f* combines WSR, ESR, and TI into a single index, offering a comprehensive measure of thrombotic risk. Since each parameter influences different aspects of thrombosis potential, *f* provides a holistic assessment of flow-mediated risk. The clear distinction in *f* values between optimal and high-risk geometries highlights its effectiveness in identifying configurations that minimize adverse flow conditions.

The parameter λ classifies flow based on its extensional and rotational quality and has been used in recent studies to assess VWF behavior^48^. It’s noted that λ magnitude does not have the same implications as other parameters, as it’s primarily descriptive of the amount of extension relative to rotation in the flow, without indicating if there is sufficient fluid stress to cause VWF extension or to trigger platelet activation. Positive λ values, when aligned with high WSR or ESR, would indicate a likelihood for VWF extension, while negative λ suggests a more rotational flow environment, where VWF tumbling is thought to maintain VWF in a globular form, leaving key platelet binding domains hidden. Given its current application primarily in numerical modeling and limited experimental validation, we prioritize WSR, ESR, and TI as the primary hemodynamic metrics in this study, evaluated together through *f*.

Current antithrombotic strategies focus on biochemical pathways but have limited efficacy mechanical pathways driving thrombosis^16,26,49–51^. Traditional anti-platelet treatments like aspirin have minimal effect on SIPA, while anticoagulants target the coagulation cascade, which incorporates multiple reactions that require low convection/flow that won’t wash away reaction products^52^. This may explain the limited efficacy of current antithrombotic strategies in preventing mBTTS thrombosis. Caplacizumab, the only drug targeting mechanical pathways, indirectly inhibits binding of VWF to GPIb-IX-V through stabilization of the autoinhibitory module that otherwise moves through force, but this drug remains expensive and has typically been used for thrombotic thrombocytopenia purpura, but not for reducing thrombus risk in surgery^53^. Our findings suggest that geometric optimization, aimed at minimizing adverse flow characteristics, could complement pharmacological strategies, potentially reducing thrombosis risk without increasing bleeding risks associated with antithrombotics.

### Clinical Translation and Surgical Implications

Our computational findings suggest specific surgical adjustments to improve mBTTS outcomes. These include placing the shunt distally relative to the right carotid artery bifurcation and using a 60° insertion angle into the pulmonary artery. While patient anatomical variations (e.g., vessel size and branching) may limit full implementation, these hemodynamically informed engineering principles should guide shunt size and placement decisions. In some cases, these adjustments can be made without added complexity, optimizing outcomes while maintaining the core surgical approach.

The 8–12% rate of mBTTS thrombosis, with 50% mortality upon occlusion, underscores the need for better strategies^1–7^. Our findings suggest that smaller pulmonary arteries (≤3.75 mm) paired with larger shunt diameters (4.0 mm) improve flow, challenging traditional views on vessel size matching. However, these recommendations must be weighed against individualized anatomical constraints and surgical risks. Notably, oversized shunts can steal flow from coronary and systemic circulation, risking cardiac arrest even without thrombosis.

In some patients, these findings could be implemented with minimal changes to current surgical practice. Pre-operative planning should consider shunt placement relative to the right carotid artery, and intraoperative consideration should be placed on maintaining acute insertion angles. Shunt-to-vessel diameter ratio also warrants attention due to its impact on flow. While exact values may vary, general trends, like favoring acute over obtuse angles, are more important. Further studies are needed to fully define the geometric parameter space.

While these theoretical recommendations may not be applicable to every patient, they could help identify those at higher risk for thrombosis, guiding clinical vigilance and management. Geometric optimization complements pharmacologic therapy by targeting mechanical contributors to clot formation without requiring new drugs or protocols. These findings also underscore the value of patient-specific preoperative modeling, enabling surgeons to tailor shunt geometry to individual anatomy.

### Limitations and Future Directions

This study has several limitations. The geometric features were derived from a retrospective cohort with a small sample size, limiting generalizability. While evaluating 54 configurations improves statistical strength, and two cardiac imaging specialists independently reviewed the imaging to reduce interpretation errors, retrospective design remains an inherent constraint.

From a computational standpoint, the model included several simplifications. We used steady-state inlet conditions, neglecting pulsatile flow, and modeled blood as a Newtonian fluid, which may underestimate effects at low shear rates. While mesh independence was confirmed, resolution limits may miss fine-scale gradients at sharp transitions. The k-ω turbulence model with low-Reynolds corrections may not fully capture transitional flows. Assumptions of rigid vessel walls and simplified boundary conditions excluded compliance and distal vascular bed resistances. Lastly, only a subset of geometric variables was analyzed, leaving others unexplored.

Validation with *in vitro* flow models is needed to confirm computational predictions. Patient-specific simulations that include full anatomy and pulsatile flow could refine geometric recommendations. Large animal studies may help assess clinical impact prior to clinical trials. Future development of real-time surgical guidance tools could support intraoperative application of these findings. Ultimately, combining geometric optimization with novel antithrombotic strategies may offer a comprehensive approach to reducing mBTTS thrombosis risk.

## CONCLUSIONS

CFD analysis of the mBTTS identified specific geometric configurations that may reduce thrombosis risk. Three key findings emerged: First, distal shunt placement relative to the right carotid artery consistently lowered prothrombotic flow; second, an acute (60°) insertion angle into the pulmonary artery reduced adverse flow at bifurcations; and third, smaller pulmonary artery diameters (≤3.75 mm) paired with larger shunt diameters (4.0 mm) produced the most favorable flow profiles, though these must be balanced against patient-specific anatomy. The optimal configuration (combining distal placement, acute angle, and appropriate vessel sizing) reduced peak adverse flow parameters by ∼50% compared to less favorable geometries. Even if some of these features can be adjusted during surgery, the findings offer a practical opportunity to optimize shunt design using an evidence-based, hemodynamics-informed approach.

Our findings challenge traditional surgical approaches, which prioritize anatomical accessibility over geometric optimization to reduce thrombogenic flow. The strong relationship between shunt geometry and hemodynamic parameters suggests that even modest adjustments to surgical technique could meaningfully impact post-operative outcomes, provided that future work validates these computational predictions. This work lays the groundwork for surgical guidelines that directly address mechanical contributors to thrombosis in mBTTS and similar procedures. Integrating geometry-based optimization with current antithrombotic strategies may offer a complementary and impactful approach to improving outcomes in this high-risk population.

## Data Availability

All data produced in the present study are available upon reasonable request to the authors

---

**Supplemental Figure E1.**
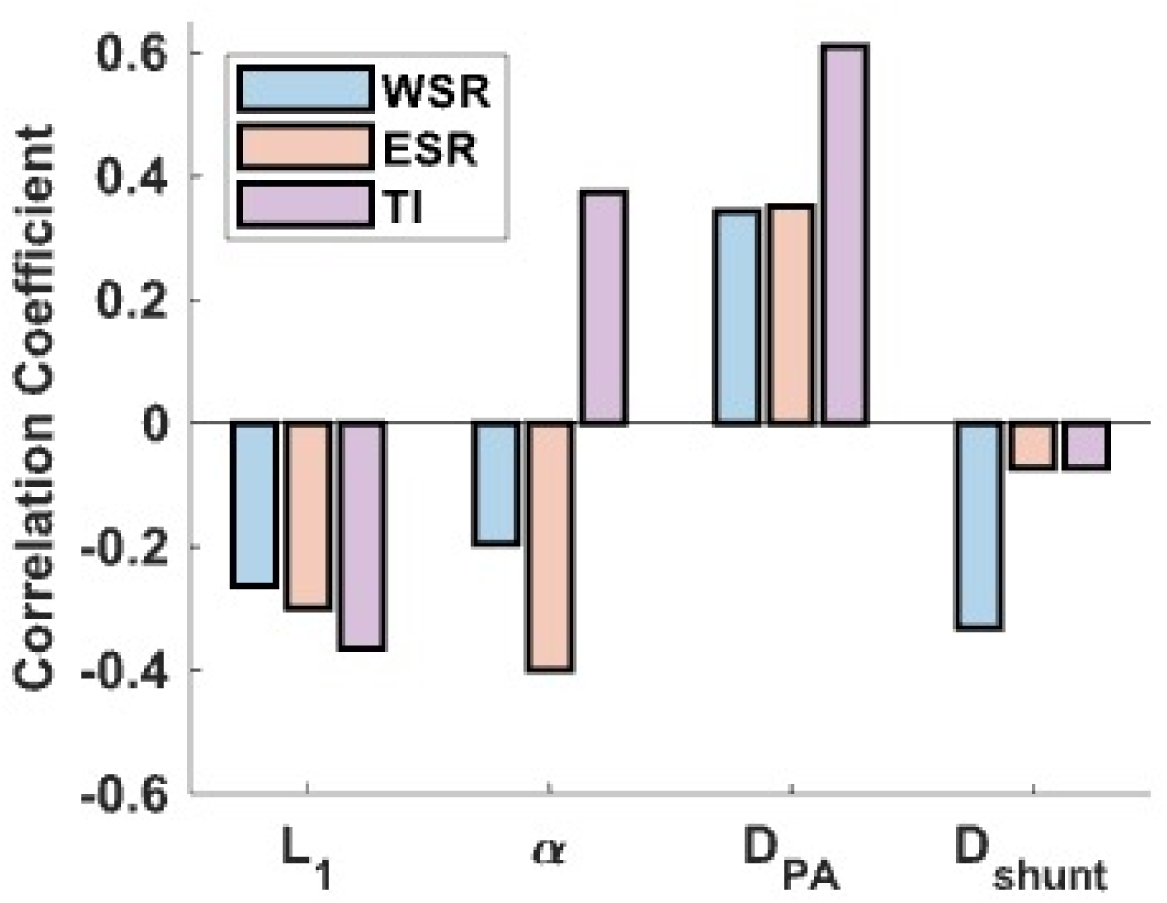
Correlation Between Geometric Parameters and Flow Characteristics. Spearman correlation coefficients (r) across 54 mBTTS configurations, showing strength and direction of associations between geometric features and flow metrics. Positive (upward) and negative (downward) bars indicate direct and inverse relationships, respectively.

**Supplemental Figure E2.**
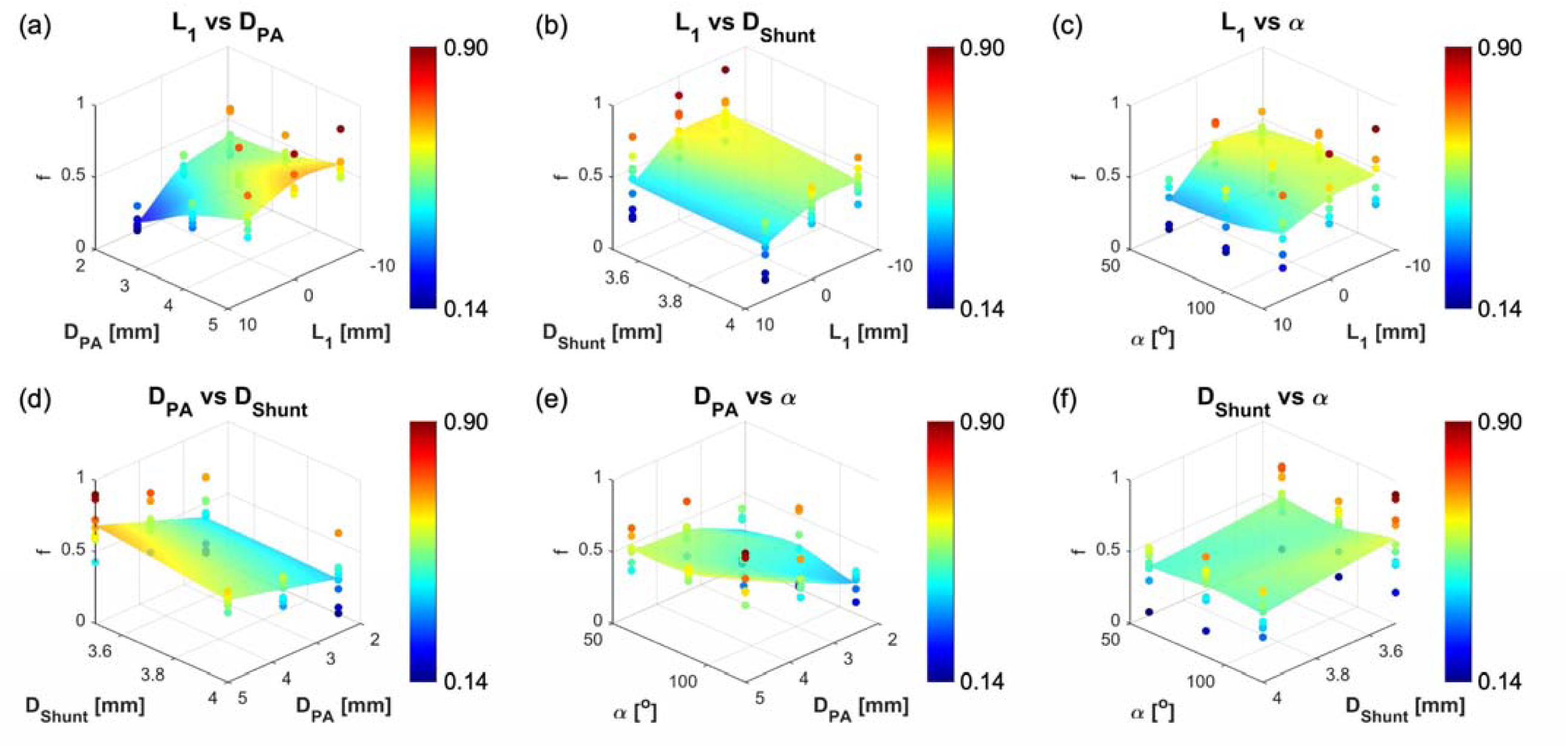
Distribution of Optimization Factor *f* Across Geometric Parameter Pairs. Surface plots showing interactions between parameter pairs: (a) L_l_ vs. D_PA_ (b) L_l_ vs. D_shunt_, (c) L_l_ vs. α, (d) D_PA_ vs. D_shunt_, (e) D_PA_ vs. α, and (f) D_shunt_ vs. α. Optimization factor *f* (range: 0.14– 0.90) reflects flow performance based on WSR, ESR, and TI, with lower *f* indicating more favorable flow.

**Supplemental Table E1:** Vessel Diameters and Flow Ratios by Pulmonary Artery Size. Vessel diameters and Murray’s Law-based outflow ratios for three (D_PA_) configurations, showing physiologically appropriate flow splitting at bifurcations. RCA: Right Carotid Artery; RSCA: Right Subclavian Artery; LPA: Left Pulmonary Artery; RPA: Right Pulmonary Artery.

|  | **RCA** | **RSCA** | **LPA** | **RPA** |
| --- | --- | --- | --- | --- |
| **$D_{PA} = 2.5$ mm** |  |  |  |  |
| Diameter (mm) | 2.6 | 3.08 | 2.5 | 2.5 |
| Outflow Ratio (%) | 22.5 | 37.5 | 20 | 20 |
| **$D_{PA} = 3.75$ mm** |  |  |  |  |
| Diameter (mm) | 2.6 | 3.08 | 3.75 | 3.75 |
| Outflow Ratio (%) | 11.6 | 19.2 | 34.6 | 34.6 |
| **$D_{PA} = 5.0$ mm** |  |  |  |  |
| Diameter (mm) | 2.6 | 3.08 | 5.0 | 5.0 |
| Outflow Ratio (%) | 5.9 | 9.9 | 42.1 | 42.1 |

**Supplemental Table E2:**
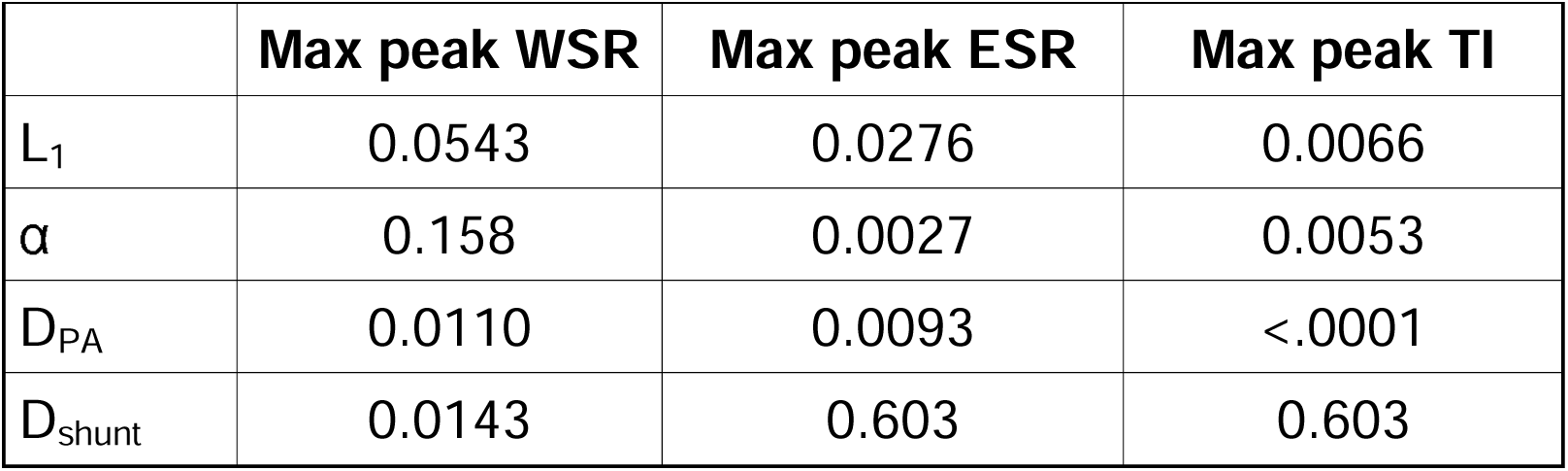
Statistical Significance of Geometric Influence on Flow Metrics. *p*-values testing correlations between geometric parameters and flow characteristics (WSR, ESR, TI). Significant values (*p* < 0.05) indicate strong evidence that a parameter influences the corresponding metric.

|  | Max peak WSR | Max peak ESR | Max peak TI |
| --- | --- | --- | --- |
| $L_1$ | 0.0543 | 0.0276 | 0.0066 |
| $\alpha$ | 0.158 | 0.0027 | 0.0053 |
| $D_{PA}$ | 0.0110 | 0.0093 | <.0001 |
| $D_{shunt}$ | 0.0143 | 0.603 | 0.603 |

**Table E3:**
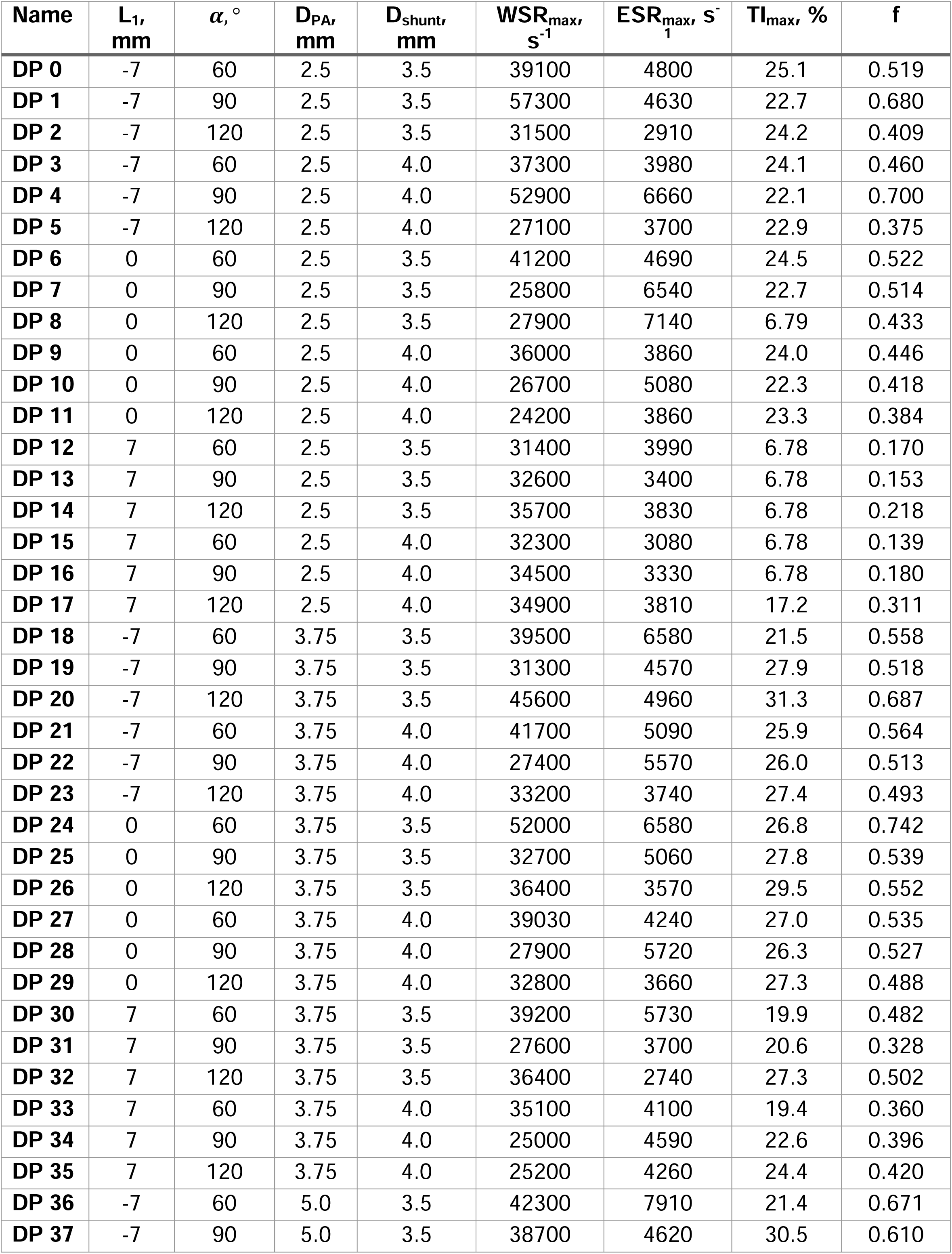
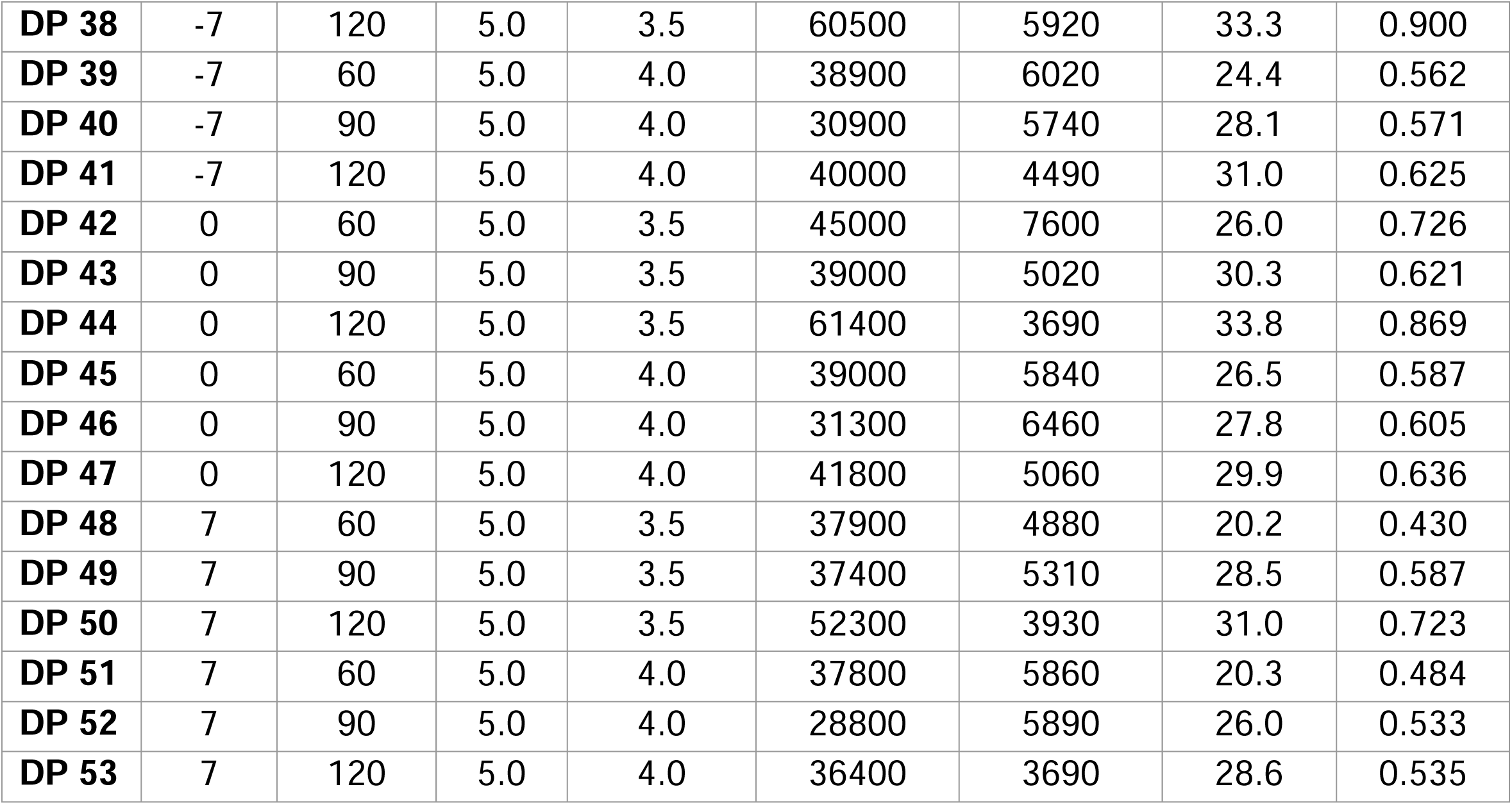
Maximal generated Wall Shear Rate (WSR), Elongational Strain Rate (ESR), and Turbulence intensity (TI) for each of the 54 corresponding geometric configurations.

| Name | L<sub>1</sub>,<br>mm | $\alpha$, ° | D<sub>PA</sub>,<br>mm | D<sub>shunt</sub>,<br>mm | WSR<sub>max</sub>,<br>s<sup>-1</sup> | ESR<sub>max</sub>, s<sup>-1</sup> | TI<sub>max</sub>, % | f |
| --- | --- | --- | --- | --- | --- | --- | --- | --- |
| DP 0 | -7 | 60 | 2.5 | 3.5 | 39100 | 4800 | 25.1 | 0.519 |
| DP 1 | -7 | 90 | 2.5 | 3.5 | 57300 | 4630 | 22.7 | 0.680 |
| DP 2 | -7 | 120 | 2.5 | 3.5 | 31500 | 2910 | 24.2 | 0.409 |
| DP 3 | -7 | 60 | 2.5 | 4.0 | 37300 | 3980 | 24.1 | 0.460 |
| DP 4 | -7 | 90 | 2.5 | 4.0 | 52900 | 6660 | 22.1 | 0.700 |
| DP 5 | -7 | 120 | 2.5 | 4.0 | 27100 | 3700 | 22.9 | 0.375 |
| DP 6 | 0 | 60 | 2.5 | 3.5 | 41200 | 4690 | 24.5 | 0.522 |
| DP 7 | 0 | 90 | 2.5 | 3.5 | 25800 | 6540 | 22.7 | 0.514 |
| DP 8 | 0 | 120 | 2.5 | 3.5 | 27900 | 7140 | 6.79 | 0.433 |
| DP 9 | 0 | 60 | 2.5 | 4.0 | 36000 | 3860 | 24.0 | 0.446 |
| DP 10 | 0 | 90 | 2.5 | 4.0 | 26700 | 5080 | 22.3 | 0.418 |
| DP 11 | 0 | 120 | 2.5 | 4.0 | 24200 | 3860 | 23.3 | 0.384 |
| DP 12 | 7 | 60 | 2.5 | 3.5 | 31400 | 3990 | 6.78 | 0.170 |
| DP 13 | 7 | 90 | 2.5 | 3.5 | 32600 | 3400 | 6.78 | 0.153 |
| DP 14 | 7 | 120 | 2.5 | 3.5 | 35700 | 3830 | 6.78 | 0.218 |
| DP 15 | 7 | 60 | 2.5 | 4.0 | 32300 | 3080 | 6.78 | 0.139 |
| DP 16 | 7 | 90 | 2.5 | 4.0 | 34500 | 3330 | 6.78 | 0.180 |
| DP 17 | 7 | 120 | 2.5 | 4.0 | 34900 | 3810 | 17.2 | 0.311 |
| DP 18 | -7 | 60 | 3.75 | 3.5 | 39500 | 6580 | 21.5 | 0.558 |
| DP 19 | -7 | 90 | 3.75 | 3.5 | 31300 | 4570 | 27.9 | 0.518 |
| DP 20 | -7 | 120 | 3.75 | 3.5 | 45600 | 4960 | 31.3 | 0.687 |
| DP 21 | -7 | 60 | 3.75 | 4.0 | 41700 | 5090 | 25.9 | 0.564 |
| DP 22 | -7 | 90 | 3.75 | 4.0 | 27400 | 5570 | 26.0 | 0.513 |
| DP 23 | -7 | 120 | 3.75 | 4.0 | 33200 | 3740 | 27.4 | 0.493 |
| DP 24 | 0 | 60 | 3.75 | 3.5 | 52000 | 6580 | 26.8 | 0.742 |
| DP 25 | 0 | 90 | 3.75 | 3.5 | 32700 | 5060 | 27.8 | 0.539 |
| DP 26 | 0 | 120 | 3.75 | 3.5 | 36400 | 3570 | 29.5 | 0.552 |
| DP 27 | 0 | 60 | 3.75 | 4.0 | 39030 | 4240 | 27.0 | 0.535 |
| DP 28 | 0 | 90 | 3.75 | 4.0 | 27900 | 5720 | 26.3 | 0.527 |
| DP 29 | 0 | 120 | 3.75 | 4.0 | 32800 | 3660 | 27.3 | 0.488 |
| DP 30 | 7 | 60 | 3.75 | 3.5 | 39200 | 5730 | 19.9 | 0.482 |
| DP 31 | 7 | 90 | 3.75 | 3.5 | 27600 | 3700 | 20.6 | 0.328 |
| DP 32 | 7 | 120 | 3.75 | 3.5 | 36400 | 2740 | 27.3 | 0.502 |
| DP 33 | 7 | 60 | 3.75 | 4.0 | 35100 | 4100 | 19.4 | 0.360 |
| DP 34 | 7 | 90 | 3.75 | 4.0 | 25000 | 4590 | 22.6 | 0.396 |
| DP 35 | 7 | 120 | 3.75 | 4.0 | 25200 | 4260 | 24.4 | 0.420 |
| DP 36 | -7 | 60 | 5.0 | 3.5 | 42300 | 7910 | 21.4 | 0.671 |
| DP 37 | -7 | 90 | 5.0 | 3.5 | 38700 | 4620 | 30.5 | 0.610 |
| **DP 38** | -7 | 120 | 5.0 | 3.5 | 60500 | 5920 | 33.3 | 0.900 |
| **DP 39** | -7 | 60 | 5.0 | 4.0 | 38900 | 6020 | 24.4 | 0.562 |
| **DP 40** | -7 | 90 | 5.0 | 4.0 | 30900 | 5740 | 28.1 | 0.571 |
| **DP 41** | -7 | 120 | 5.0 | 4.0 | 40000 | 4490 | 31.0 | 0.625 |
| **DP 42** | 0 | 60 | 5.0 | 3.5 | 45000 | 7600 | 26.0 | 0.726 |
| **DP 43** | 0 | 90 | 5.0 | 3.5 | 39000 | 5020 | 30.3 | 0.621 |
| **DP 44** | 0 | 120 | 5.0 | 3.5 | 61400 | 3690 | 33.8 | 0.869 |
| **DP 45** | 0 | 60 | 5.0 | 4.0 | 39000 | 5840 | 26.5 | 0.587 |
| **DP 46** | 0 | 90 | 5.0 | 4.0 | 31300 | 6460 | 27.8 | 0.605 |
| **DP 47** | 0 | 120 | 5.0 | 4.0 | 41800 | 5060 | 29.9 | 0.636 |
| **DP 48** | 7 | 60 | 5.0 | 3.5 | 37900 | 4880 | 20.2 | 0.430 |
| **DP 49** | 7 | 90 | 5.0 | 3.5 | 37400 | 5310 | 28.5 | 0.587 |
| **DP 50** | 7 | 120 | 5.0 | 3.5 | 52300 | 3930 | 31.0 | 0.723 |
| **DP 51** | 7 | 60 | 5.0 | 4.0 | 37800 | 5860 | 20.3 | 0.484 |
| **DP 52** | 7 | 90 | 5.0 | 4.0 | 28800 | 5890 | 26.0 | 0.533 |
| **DP 53** | 7 | 120 | 5.0 | 4.0 | 36400 | 3690 | 28.6 | 0.535 |

## Glossary of Abbreviations

CHD: Congenital Heart Disease
mBTTS: modified Blalock-Taussig-Thomas shunt
SIPA: shear induced platelet activation & aggregation
VWF: von Willebrand Factor
CT: computer tomography
MR: magnetic resonance
HLHS: hypoplastic left heart syndrome
PA: pulmonary artery
STS: society of thoracic surgeons
CFD: computational fluid dynamics
WSR: wall shear rate
ESR: elongational strain rate
TI: turbulence intensity.
Re: Reynolds number

---

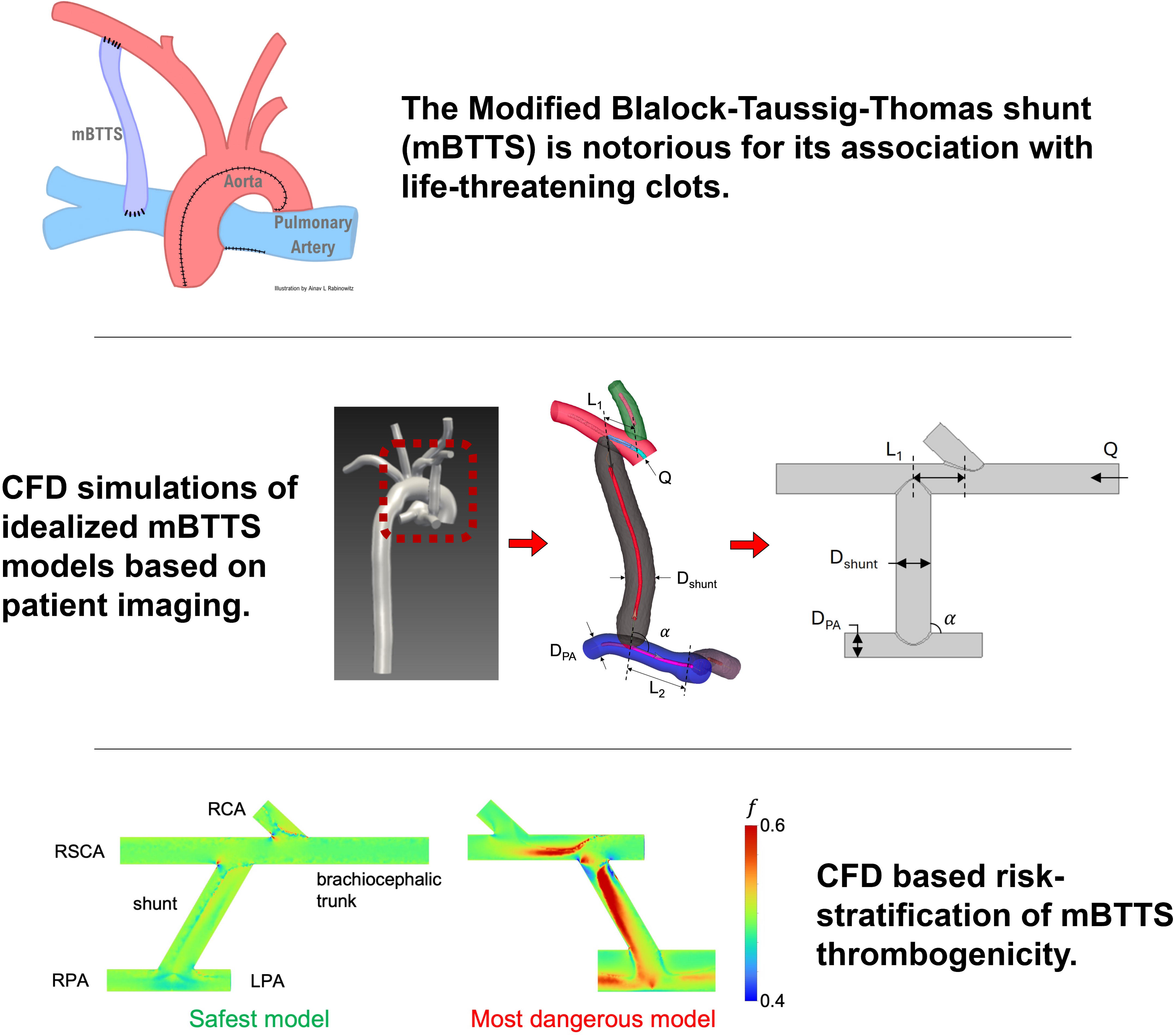

## REFERENCES

1. Giglia TM, Massicotte MP, Tweddell JS, et al. Prevention and treatment of thrombosis in pediatric and congenital heart disease: a scientific statement from the American Heart Association. Circulation. Dec 17 2013;128(24):2622–703. doi:10.1161/01.cir.0000436140.77832.7a

2. Penk JS, Reddy S, Polito A, et al. Bleeding and Thrombosis With Pediatric Extracorporeal Life Support: A Roadmap for Management, Research, and the Future From the Pediatric Cardiac Intensive Care Society: Part 2. Pediatr Crit Care Med. Nov 2019;20(11):1034-1039. doi:10.1097/pcc.0000000000002104

3. Penk JS, Reddy S, Polito A, et al. Bleeding and Thrombosis With Pediatric Extracorporeal Life Support: A Roadmap for Management, Research, and the Future From the Pediatric Cardiac Intensive Care Society: Part 1. Pediatr Crit Care Med. Nov 2019;20(11):1027-1033. doi:10.1097/pcc.0000000000002054

4. Giglia TM, Witmer C. Bleeding and Thrombosis in Pediatric Cardiac Intensive Care. Pediatr Crit Care Med. Aug 2016;17(8 Suppl 1):S287-95. doi:10.1097/pcc.0000000000000814

5. Monagle P. Thrombosis: congenital heart disease and thrombosis: what do we know? Nat Rev Cardiol. Mar 2014;11(3):132–4. doi:10.1038/nrcardio.2013.217

6. Garven E, Rodell CB, Shema K, et al. Tunable Blood Shunt for Neonates With Complex Congenital Heart Defects. Original Research. Frontiers in Bioengineering and Biotechnology. 2022-January-13 2022;9doi:10.3389/fbioe.2021.734310

7. Petrucci O, O’Brien SM, Jacobs ML, Jacobs JP, Manning PB, Eghtesady P. Risk factors for mortality and morbidity after the neonatal Blalock-Taussig shunt procedure. Ann Thorac Surg. Aug 2011;92(2):642–51; discussion 651-2. doi:10.1016/j.athoracsur.2011.02.030

8. Al Jubair KA, Al Fagih MR, Al Jarallah AS, et al. Results of 546 Blalock-Taussig shunts performed in 478 patients. Cardiol Young. Oct 1998;8(4):486–90. doi:10.1017/s1047951100007150

9. Fenton KN, Siewers RD, Rebovich B, Pigula FA. Interim mortality in infants with systemic-to-pulmonary artery shunts. Ann Thorac Surg. Jul 2003;76(1):152–6; discussion 156-7. doi:10.1016/s0003-4975(03)00168-1

10. Chow TW, Hellums JD, Moake JL, Kroll MH. Shear stress-induced von Willebrand factor binding to platelet glycoprotein Ib initiates calcium influx associated with aggregation. Blood. Jul 1 1992;80(1):113–20.

11. Hellums JD. 1993 Whitaker Lecture: biorheology in thrombosis research. Ann Biomed Eng. Sep-Oct 1994;22(5):445-55. doi:10.1007/bf02367081

12. Susen S, Rauch A, Van Belle E, Vincentelli A, Lenting PJ. Circulatory support devices: fundamental aspects and clinical management of bleeding and thrombosis. J Thromb Haemost. Oct 2015;13(10):1757–67. doi:10.1111/jth.13120

13. Uriel N, Pak SW, Jorde UP, et al. Acquired von Willebrand syndrome after continuous-flow mechanical device support contributes to a high prevalence of bleeding during long-term support and at the time of transplantation. J Am Coll Cardiol. Oct 5 2010;56(15):1207–13. doi:10.1016/j.jacc.2010.05.016

14. Penn E, Qiao Y, Platten K, et al. Impact of Vascular Geometry on Thrombosis in Pediatric Patients With Modified Blalock-Taussig-Thomas Shunt: A Pilot Study. Journal of the American Heart Association. 2025;14(6)

15. Sing CE, Alexander-Katz A. Elongational flow induces the unfolding of von Willebrand factor at physiological flow rates. Biophysical Journal. 2010;98(9):L35–L37. doi:10.1016/j.bpj.2010.01.032

16. Bark Jr. DL, Para AN, Ku DN. Correlation of thrombosis growth rate to pathological wall shear rate during platelet accumulation. Biotechnology and Bioengineering. 2012;109(10):2642–2650. 10.1002/bit.24537

17. Nesbitt WS, Westein E, Tovar-Lopez FJ, et al. A shear gradient-dependent platelet aggregation mechanism drives thrombus formation. Nat Med. Jun 2009;15(6):665-73. doi:10.1038/nm.1955

18. Shankaran H, Alexandridis P, Neelamegham S. Aspects of hydrodynamic shear regulating shear-induced platelet activation and self-association of von Willebrand factor in suspension. Blood. Apr 1 2003;101(7):2637–45. doi:10.1182/blood-2002-05-1550

19. Moake JL, Turner NA, Stathopoulos NA, Nolasco LH, Hellums JD. Involvement of large plasma von Willebrand factor (vWF) multimers and unusually large vWF forms derived from endothelial cells in shear stress-induced platelet aggregation. J Clin Invest. Dec 1986;78(6):1456–61. doi:10.1172/jci112736

20. Zhang JN, Bergeron AL, Yu Q, et al. Platelet aggregation and activation under complex patterns of shear stress. Thromb Haemost. Nov 2002;88(5):817–21.

21. Nobili M, Sheriff J, Morbiducci U, Redaelli A, Bluestein D. Platelet activation due to hemodynamic shear stresses: damage accumulation model and comparison to in vitro measurements. Asaio j. Jan-Feb 2008;54(1):64–72. doi:10.1097/MAT.0b013e31815d6898

22. Herbig BA, Diamond SL. Pathological von Willebrand factor fibers resist tissue plasminogen activator and ADAMTS13 while promoting the contact pathway and shear-induced platelet activation. J Thromb Haemost. 2015;13(9):1699–1708. doi:10.1111/jth.13044

23. Springer TA. von Willebrand factor, Jedi knight of the bloodstream. Blood. 2014;124(9):1412–1425. doi:10.1182/blood-2014-05-378638

24. Bozzi S, Dominissini D, Redaelli A, Passoni G. The effect of turbulence modelling on the assessment of platelet activation. J Biomech. Nov 9 2021;128:110704. doi:10.1016/j.jbiomech.2021.110704

25. Stein PD, Sabbah HN. Measured turbulence and its effect on thrombus formation. Circ Res. Oct 1974;35(4):608–14. doi:10.1161/01.res.35.4.608

26. Grande Gutiérrez N, Mukherjee D, Bark D, Jr. Decoding thrombosis through code: a review of computational models. J Thromb Haemost. Aug 30 2023;doi:10.1016/j.jtha.2023.08.021

27. Williams D, Leuthardt EC, Genin GM, Zayed M. Tailoring of arteriovenous graft-to-vein anastomosis angle to attenuate pathological flow fields. Sci Rep. Jun 9 2021;11(1):12153. doi:10.1038/s41598-021-90813-3

28. Buehler MJ, Genin GM. Integrated multiscale biomaterials experiment and modelling: a perspective. Interface Focus. Feb 6 2016;6(1):20150098. doi:10.1098/rsfs.2015.0098

29. Kong F, Stocker S, Choi PS, Ma M, Ennis DB, Marsden AL. SDF4CHD: Generative modeling of cardiac anatomies with congenital heart defects. Med Image Anal. Oct 2024;97:103293. doi:10.1016/j.media.2024.103293

30. Kong F, Shadden SC. Learning Whole Heart Mesh Generation From Patient Images for Computational Simulations. IEEE Trans Med Imaging. Feb 2023;42(2):533–545. doi:10.1109/tmi.2022.3219284

31. Kong F, Shadden SC. Automating Model Generation for Image-Based Cardiac Flow Simulation. J Biomech Eng. Nov 1 2020;142(11)doi:10.1115/1.4048032

32. Murray CD. The Physiological Principle of Minimum Work: I. The Vascular System and the Cost of Blood Volume. Proc Natl Acad Sci U S A. Mar 1926;12(3):207–14. doi:10.1073/pnas.12.3.207

33. Astarita G. Objective and generally applicable criteria for flow classification. Journal of Non-Newtonian Fluid Mechanics. 1979;6(1):69–76.

34. Ahmad U, Fatimi SH, Naqvi I, et al. Modified Blalock-Taussig shunt: immediate and short-term follow-up results in neonates. Heart Lung Circ. Feb 2008;17(1):54–8. doi:10.1016/j.hlc.2007.06.003

35. de Leval MR, McKay R, Jones M, Stark J, Macartney FJ. Modified Blalock-Taussig shunt. Use of subclavian artery orifice as flow regulator in prosthetic systemic-pulmonary artery shunts. J Thorac Cardiovasc Surg. Jan 1981;81(1):112–9.

36. Ilbawi MN, Grieco J, DeLeon SY, et al. Modified Blalock-Taussig shunt in newborn infants. J Thorac Cardiovasc Surg. Nov 1984;88(5 Pt 1):770-5.

37. O’Connor MJ, Ravishankar C, Ballweg JA, et al. Early systemic-to-pulmonary artery shunt intervention in neonates with congenital heart disease. J Thorac Cardiovasc Surg. Jul 2011;142(1):106–12. doi:10.1016/j.jtcvs.2010.10.033

38. Wells WJ, Yu RJ, Batra AS, Monforte H, Sintek C, Starnes VA. Obstruction in modified Blalock shunts: a quantitative analysis with clinical correlation. Ann Thorac Surg. Jun 2005;79(6):2072–6. doi:10.1016/j.athoracsur.2004.12.050

39. Diane M Pickles and Kirstie Keller P. The Cost of Complex Congenital Heart Disease in the US. 2024. https://www.additionalventures.org/wp-content/uploads/2024/04/The-Cost-of-cCHD-in-the-US-Additional-Ventures-2024-04-05.pdf

40. Rabinowitz EJ, Ouyang A, Armstrong DR, Wallendorf M, Said AS. Poor Reliability of Common Measures of Anticoagulation in Pediatric Extracorporeal Membrane Oxygenation. Asaio j. Jun 1 2022;68(6):850–858. doi:10.1097/mat.0000000000001582

41. Hellums JD. 1993 Whitaker Lecture: biorheology in thrombosis research. Ann Biomed Eng. 1994;22(5):445-455.

42. Chow TW, Hellums JD, Moake JL, Kroll MH. Shear stress-induced von Willebrand factor binding to platelet glycoprotein Ib initiates calcium influx associated with aggregation. 1992;

43. Susen S, Rauch A, Van Belle E, Vincentelli A, Lenting P. Circulatory support devices: fundamental aspects and clinical management of bleeding and thrombosis. J Thromb Haemost. 2015;13(10):1757–1767.

44. Uriel N, Pak S-W, Jorde UP, et al. Acquired von Willebrand syndrome after continuous-flow mechanical device support contributes to a high prevalence of bleeding during long-term support and at the time of transplantation. Journal of the American College of Cardiology. 2010;56(15):1207–1213.

45. Deng W, Xu Y, Chen W, et al. Platelet clearance via shear-induced unfolding of a membrane mechanoreceptor. Nature Communications; 2016.

46. Sheriff J, Soares JS, Xenos M, Jesty J, Bluestein D. Evaluation of shear-induced platelet activation models under constant and dynamic shear stress loading conditions relevant to devices. 2013;41(6):1279–1296.

47. Bortot M, Ashworth K, Sharifi A, et al. Turbulent Flow Promotes Cleavage of VWF (von Willebrand Factor) by ADAMTS13 (A Disintegrin and Metalloproteinase With a Thrombospondin Type-1 Motif, Member 13). 2019;39(9):1831-1842. doi:10.1161/ATVBAHA.119.312814

48. Zhussupbekov M, Rojano RM, Wu W-T, Antaki JF. von Willebrand factor unfolding mediates platelet deposition in a model of high-shear thrombosis. 2022;121(21):4033–4047.

49. Bark DL, Jr., Ku DN. Wall shear over high degree stenoses pertinent to atherothrombosis. J Biomech. Nov 16 2010;43(15):2970–7. doi:10.1016/j.jbiomech.2010.07.011

50. Bark DL, Jr., Ku DN. Platelet transport rates and binding kinetics at high shear over a thrombus. Biophys J. Jul 16 2013;105(2):502–11. doi:10.1016/j.bpj.2013.05.049

51. Sharifi A, Bark D. Mechanical Forces Impacting Cleavage of Von Willebrand Factor in Laminar and Turbulent Blood Flow. Fluids. 2021;6(2):67.

52. Turner NA, Moake JL, Kamat SG, et al. Comparative real-time effects on platelet adhesion and aggregation under flowing conditions of in vivo aspirin, heparin, and monoclonal antibody fragment against glycoprotein IIb-IIIa. Circulation. Mar 1 1995;91(5):1354-62. doi:10.1161/01.cir.91.5.1354

53. Arce NA, Cao W, Brown AK, et al. Activation of von Willebrand factor via mechanical unfolding of its discontinuous autoinhibitory module. Nature Communications. 2021;12(1)(2360)doi:10.1038/s41467-021-22634-x

